# Automating Responses to Patient Portal Messages Using Generative AI

**DOI:** 10.1101/2024.04.25.24306183

**Authors:** Amarpreet Kaur, Alex Budko, Katrina Liu, Eric Eaton, Bryan Steitz, Kevin B. Johnson

## Abstract

**Background:** Patient portals serve as vital bridges between patients and providers, playing an increasing role in healthcare communication. The rising volume and complexity of these messages is exacerbating physician and nursing burnout. Recent studies have demonstrated that AI chatbots can generate message responses that are viewed favorably by healthcare professionals; however, these studies have not included the diverse range of messages typically found in patient portals. Our goal is to investigate the quality of GPT-generated message responses across the spectrum of message types within a patient portal.

**Methods:** We used novel prompt engineering techniques to craft synthetic responses tailored to adult primary care patients. We enrolled a sample of primary care providers in a cross-sectional study to compare authentic with synthetic patient portal message responses, generated by GPT-4. The survey assessed each message’s empathy, relevance, medical accuracy, and readability on a scale from 0 to 5. Respondents were asked to identify messages that were GPT-generated vs. provider-generated. Mean scores for all metrics were computed for subsequent analysis.

**Results:** A total of 49 health care providers participated in the survey (59% completion rate), comprising 16 physicians and 32 advanced practice providers (APPs). When presented with GPT vs. authentic message response pairs, participants correctly identified GPT-generated responses 73% of the time and correctly identified authentic responses 50% of the time. In comparison to messages generated by physicians, GPT-4 generated messages exhibited higher mean scores for empathy (3.57 vs. 3.07, p < 0.001), relevance (3.94 vs. 3.81, p = 0.08) accuracy (4.05 vs. 3.95, p= 0.12) and readability (4.5 vs. 4.13, p < 0.001).

**Limitations:** The study is a single site, single-specialty study, limited due to the use of synthetic data.

**Conclusion:** Our findings affirm the potential of GPT-generated patient portal message responses to achieve comparable levels of empathy, relevance, and readability to those found in typical responses according to the health care providers and indicates promising prospects for their integration in the healthcare sector. Additional studies should be done within provider workflows and with careful evaluation of patient attitudes and concerns related to the ethics as well as the quality of generated patient portal message responses in all settings.

## INTRODUCTION

Patient portals have become an integral and indispensable component of modern healthcare, providing patients with secure online access to vital health information and facilitating crucial communication bridges between healthcare professionals (HCPs) and patients. In doing so, they foster stronger connections between providers and patients and facilitate the delivery of personalized care through effective communication.^1^ With the rapid adoption of patient portal messaging during the COVID-19 pandemic,^2^ the continually increasing volume of in-basket patient messages has emerged as a pressing issue in the healthcare sector that appears to be exacerbating health care provider burnout.^1,3–5^ First documented in 1974, physician burnout has been linked to the demands of EHR documentation, consuming substantial clinical time.^3,6^ Primary care providers face uniquely heightened burnout risks among all HCPs, emphasizing the pressing need for interventions to alleviate EHR-related burdens and support clinician well-being.^3^

Large language models, such as OpenAI^®^’s GPT-4, have emerged as a promising tool in the healthcare sector, particularly for mitigating documentation-related burnout among clinicians. GPT-4 is in a class known as generative AI—tools that use deep learning models to create content based on the data from which it was trained. These generative tasks respond to prompts created by another agent (typically a human) experienced in the careful wording necessary to align the prompt with the needs of the agent.

Generative AI has gained widespread attention in the medical community due to its capacity to effectively streamline various documentation processes, including the generation of patient clinic letters, radiology reports, medical notes, discharge summaries, and even passing the United States Medical Licensing Exam (USMLE).^7–9^ Several studies have highlighted generative AI’s effectiveness in clinical decision support.^9–11^ Recent improvements in prompt engineering through techniques such as few-shot learning have allowed automation of some in-basket message work such as patient portal message responses.^7^ Several studies have aimed to develop and evaluate the effectiveness of fine-tuned large language models (LLMs) in generating responses to patient queries.^7,12^ The objective of this study is to further explore HCP acceptability of AI-generated message responses.

## METHODS

### Initial Data Collection and Creation of Synthetic Patient Portal Message

Considering the sensitive nature of real patient portal messages, we first retrieved a set of 85 patient portal messages and clinician responses from a repository at Vanderbilt University Medical Center (VUMC). These messages were fully de-identified then manually rephrased to convey similar content but vary tone and length from the original message. Using these messages, we engineered a prompt within GPT-4 (GPT)^13^ to generate similar messages in terms of tone, length, and topic. Once the research team was satisfied with the prompt (Figure 1), we recruited a convenience sample of eight clinicians to review and compare synthetic and authentic patient portal messages. This sampling approach allowed us to use email distribution lists to contact eligible HCPs and to develop a denominator to assess completion rate. Survey analysis determined that participants correctly distinguished GPT-generated from clinician-generated messages only 51.1% of the time. Given these results, we combined our pool of messages into one set to develop our synthetic patient portal message responses. Of note, we also received de-identified responses to each message, which were not made available to the pipeline development team.

**Figure 1:**
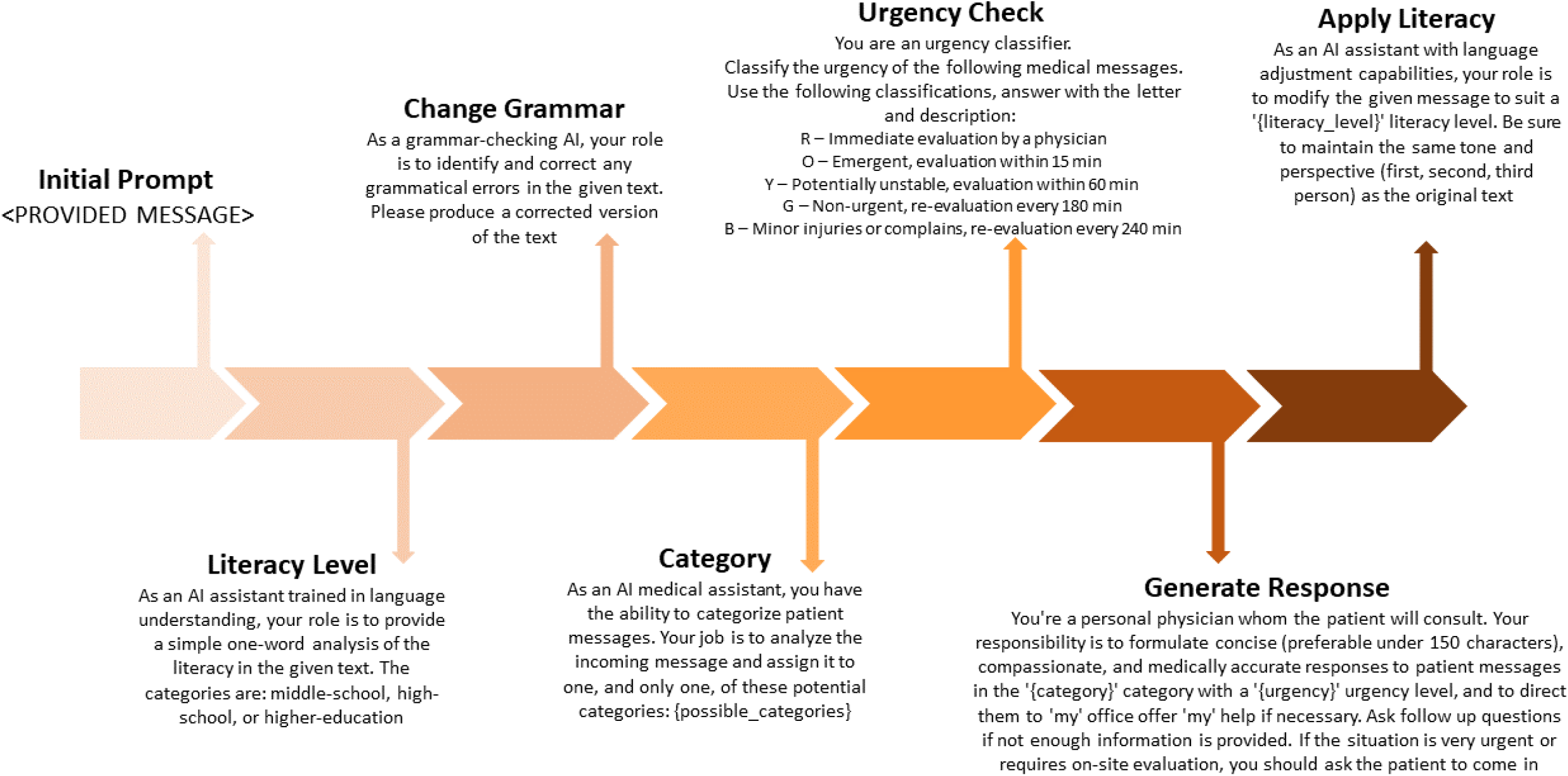
Diagram representation of the patient portal message response pipeline using the GPT-4 API

### Pipeline Development

We used GPT-4 prompt engineering without fine-tuning (known as zero-shot learning) to automate patient responses without task-specific training data. This approach leveraged the model’s pre-training knowledge to generate contextually relevant responses. However, testing revealed occasional deviations in relevance and style compared with authentic clinician responses. Therefore, we adopted a supervised learning approach with a small number of examples (known as few-shot learning) to enhance relevance. Our final engineered prompts employed a set of feature-specific prompts to refine responses. Once we were satisfied with the face validity of responses, we generated synthetic patient portal message responses across the range of categories based on work done by Cronin.^14^ The final pipeline, summarized in figure 1, tailored responses to the message’s literacy level, urgency, and context, ensuring comprehensive management of user inputs.

### Evaluation of Message Response Pairs

To evaluate the quality and authenticity of messages generated by our pipeline, we conducted a cross-sectional study of HCPs across the University of Pennsylvania Health System. We designed a survey to compare synthetic and authentic patient portal message responses and to assess the quality of each. The survey consisted of 20 questions. Each question included a synthetic patient portal message and an accompanying GPT-generated or authentic patient portal message response. For each pair, respondents were asked to rate the response according to four key quality dimensions of communication: *Empathy*, reflecting the degree of consideration for the patient’s emotions in the message; *Relevance*, assessing how closely the content addressed the patient’s expressed needs; *Medical Accuracy*, gauging the alignment of the message with established medical practices and guidelines; and *Readability*, evaluating the clarity, coherence, and simplicity of the language employed. Each quality dimension was presented as a Likert-style question with five possible responses. Additionally, participants were asked to discern whether each message response was GPT-generated or written by a real provider.

We recruited survey participants, comprising HCPs who identified as primary care MDs, DOs, and advanced practice providers (APPs), through an email distribution list. This sampling frame covered most primary care providers at our institution. Initially, information about the research project was disseminated to HCPs, inviting interested individuals to reach out to the research team via email and request access to the survey. There were 84 potential participants who responded to that request. We sent a survey link to each potential participant and upon completion of the survey, participants received a $10 Starbucks gift card as a token of appreciation. The survey was distributed using both REDCap and Google Forms, the latter being utilized due to firewall restrictions. The survey was administered between November 28, 2023, and January 5, 2024. We used Microsoft Excel (v16.83) for univariate analyses and JMP (version 17.2.0) ^17^ to analyze the impact of covariates on responses.

## RESULTS

Table 1 provides an overview of various demographic and professional variables among the 49 respondents. Most participants identified as female (77.6%), with 69% between the ages of 31 and 40. A total of 67% of respondents identified as APPs, while 33% held a medical degree (MD or DO). Years of experience seeing patients varied, with the largest group having less than five years of experience (31%), followed by experience between 10-15 years (18%). Most respondents worked in clinics (69%), in urban settings (63%), and reported receiving 25-75 in-basket messages from patients during a typical work week (55%). Most respondents (76%) indicated no or unknown experience with AI tools in medical practice.

**Table 1:** Overview of Participant Demographics, Medical Education and Specialization, and Current Medical Practices.

|  |  |
| --- | --- |
| General Demographics | n=49, n (%) |
| Gender |  |
| Male | 11 (22.4) |
| Female | 38 (77.6) |
| Age |  |
| <25 | - (0) |
| 26-30 | 5 (10.2) |
| 31-40 | 19 (38.78) |
| 41-50 | 15 (30.61) |
| 51-60 | 5 (10.2) |
| >60 | 5 (10.2) |
| Medical Degree |  |
| MD or DO | 16 (32.65) |
| Advanced Practice Provider (APP) | 33 (67.35) |
| Years of experience seeing patients |  |
| <5 | 15 (30.61) |
| 5-10 | 7 (14.29) |
| 10-15 | 9 (18.37) |
| 15-20 | 5 (10.2) |
| 20-25 | 5 (10.2) |
| 25-30 | 2 (4.08) |
| 30-35 | 4 (8.16) |
| >35 | 2 (4.08) |
| Clinical Setting |  |
| Hospital | 2 (4.08) |
| Clinic | 34 (69.39) |
| Private Setting – Solo Practice | - (0) |
| Private Setting – Group Practice with 1-5 providers | 3 (6.1) |
| Private Setting – Group Practice with >5 providers | 6 (12.24) |
| Outpatient specialty practice on hospital campus | 1 (2.04) |
| Long Term Care/ Office Split | 1 (2.04) |
| Other | 2 (4.08) |
| Geographic Location |  |
| Urban | 31 (63.27) |
| Suburban | 18 (36.73) |
| Rural | 0 (0) |
| Number of patients seen during work week |  |
| <20 | 6 (12.24) |
| 20-40 | 16 (32.65) |
| 40-60 | 11 (22.45) |
| 60-80 | 11 (22.45) |
| 80-100 | 4 (8.16) |
| >100 | 1 (2.04) |
| Number of in-Basket messages received from patients during work week |  |
| <25 | 10 (20.41) |
| 26-50 | 15 (30.61) |
| 51-75 | 12 (24.49) |
| 76-100 | 5 (10.2) |
| 101-200 | 7 (14.29) |
| >200 | - (0) |
| Experience with AI tools in medical practice |  |
| Yes | 4 (8.16) |
| No | 37 (75.51) |
| Not Sure | 8 (16.33) |

Table 2 and figure 2 summarize the overall assessment of message-response quality. Notably, GPT-generated responses generally outperformed real responses across all key characteristics, demonstrating statistical significance with empathy (p < 0.001) and readability (p < 0.001). Relevance also trended toward significance (p = 0.08). When presented with GPT vs. authentic message response pairs, participants correctly identified GPT messages 73% of the time (good guessers) and correctly identified authentic messages 50% of the time. There were no statistically significant differences between good guessers and other participants as determined by one-way ANOVA (F(1,47) = 2.27, p = 0.13).

**Figure 2:**
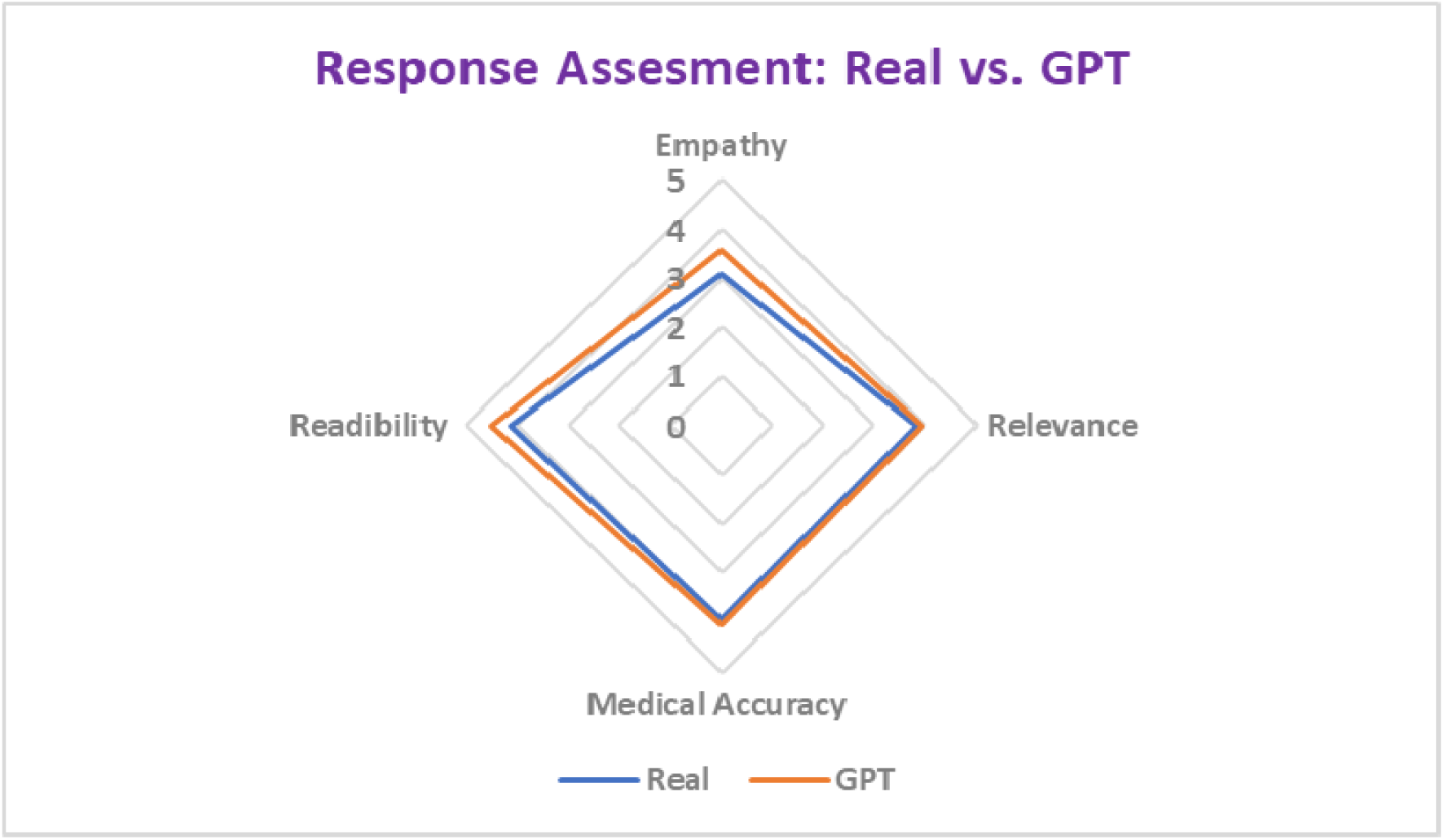
Evaluation of the Pipeline. The radar diagram illustrates the mean comparison of GPT-generated and real responses using an ordinal scale ranging from 1 (low) to 5 (high). A rating of 1 indicates poor performance, while 5 signifies excellent performance.

**Table 2:** Comparative Analysis of GPT versus real message responses. The table above provides a comprehensive breakdown of the average means and medians derived for the four key characteristics, comparing GPT-generated message-response pairs to real ones. Both empathy and readability were statistically better for GPT-generated responses.

|  | GPT Response |  | Real Response |  | Significance (t-test) |
| --- | --- | --- | --- | --- | --- |
|  | Mean (+/- SD) | Median | Mean (+/- SD) | Median | P-values |
| Empathy | 3.57 (1.02) | 3.6 | 3.07 (1.00) | 3.1 | < 0.001 |
| Relevance | 3.94 (1.00) | 4.2 | 3.81 (1.09) | 4 | 0.08 |
| Medical Accuracy | 4.05 (0.92) | 4.2 | 3.95 (0.99) | 4 | 0.12 |
| Readability | 4.50 (0.68) | 4.9 | 4.13 (1.01) | 4.7 | < 0.001 |

**Table 3:** Overview of message-response pairs along with the distribution of how participants identified these pairs. *Message category interrater agreement 95% (Cohen’s Kappa 0.94 - near perfect agreement)

| Message-Response Pair | Message Category* | Response type | % Identified as GPT (n=49) | % Identified as Real (n=49) |
| --- | --- | --- | --- | --- |
| **Message:** Ben is having a few problems. His stomach is extended with possible fluid retention. He took his fluid medicine yesterday but it doesn't seem to have helped just yet. He is also becoming very short of breath when doing anything. I wasn't sure if he could be seen in clinic soon or if he could have some blood work done to check levels.<br>**Response:** I'm really sorry to hear about Ben's symptoms. It's really important to get him checked out as soon as possible. Can you please bring him to the clinic so the doctors can give him a thorough check-up and do the right tests? | Medical Management | GPT | 36 (73%) | 13 (27%) |
| **Message:** My R elbow is hurting. If I schedule an apt, may I receive another injection? My elbow surgery is scheduled in Oct.<br>**Response:** □ I'm sorry to hear that your elbow hurts. It would be a good idea to make an appointment to have a doctor check your symptoms and talk about the possibility of getting another shot. | Medical interventions | GPT | 33 (67%) | 16 (33%) |
| **Message:** Do I need to prepare in anyway or the upcoming fluid deprivation test? such as quit drinking | Informational - | REAL | 21 (43%) | 28 (57%) |
| fluids at a certain time or avoid caffeine and alcohol in the time leading up to it?<br>**Response:** The test usually finishes faster if you minimize fluid intake after midnight. However, you do not need to strictly avoid drinking anything after midnight and should not do so if you are noticeably thirsty. | preparation |  |  |  |
| **Message:** Good morning, I have been dealing with this crud for 8 days now. I still have this terrible cough that I can not get rid of and when I breathe it doesn't feel like I am getting any air. It is keeping me up at night. I am coughing up/blowing out of my nose this bright yellow clumpy nasty stuff and I am just really tired of the cough. Any suggestions on what else I can do to help this clear up?<br>**Response:** So sorry to hear you've been dealing with this for such a long time. I'd recommend you come in for an acute visit or go to one of our walk-in clinics if closer to your home. We can be sure there isn't something that needs more than simple over the counter treatment. It's difficult to fully assess your symptoms and severity of illness without seeing you in person. | Medical - problems | REAL | 22 (45%) | 27 (55%) |
| **Message:** I need to talk to someone about possibly being referred to see a therapist. I've began to feel very unlike myself. This happened with my previous pregnancy and it led me to make some hasty not | Medical - referrals | REAL | 12 (24%) | 37 (76%) |
| very well planned decisions. ☐ I feel very defeated physically and mentally. ☐ If there is someone you can refer me to I'd appreciate it.<br>**Response:** Thank you so much for reaching out. Yes, let's get you to see a counselor so that we can get you feeling more like yourself. We have a counselor that works at our office. She is funded by a grant so your appointments with her would be free and we could schedule them the same day as your visit so that you don't have to make extra trips. Are you open to meeting with her? |  |  |  |  |
| **Message:** Please let me know when you get my results<br>**Response:** I will inform you once I receive your lab results. If you have any worries or inquiries, please don't hesitate to contact me. | Logistical - tests | GPT | 41 (84%) | 8 (16%) |
| **Message:** I am feeling much better but still have sinus pressure and congestion. I am not sure if I should start the antibiotics or not.<br>**Response:** I'm happy to hear that you're feeling better. Sometimes, even after the infection is gone, sinus pressure and congestion can still stick around. You could try using decongestants or saline nasal rinses that you can buy without a prescription. If your symptoms get worse or don't go away, please contact our office so we can check it out. | Medical - management | GPT | 21* (44%) | 27* (56%) |
| **Message:** Good morning! It looks like everything is stable and normal. So why do I feel so crummy? Will switching meds help knowing my results are normal?<br>**Response:** Your inflammation markers have not been significant elevated, so we cannot necessarily rely on them to assess your disease . Treatment change is recommended based on your report of increased symptoms of joint stiffness, swelling and improvement after taking steroids. | Medical - management | REAL | 29 (59%) | 20 (41%) |
| **Message:** Ben is having a few problems. His stomach is extended with possible fluid retention. He took his fluid medicine yesterday but it doesn't seem to have helped just yet. He is also becoming very short of breath when doing anything. I wasn't sure if he could be seen in clinic soon or if he could have some blood work done to check levels.<br>**Response:** I spoke with [DOCTOR] about this. Please continue to monitor for the next 24hrs. If his condition continues to worsen, he should go to the ER. [DOCTOR] will call you this afternoon to check in | Medical - management | REAL | 23 (47%) | 26 (53%) |
| **Message:** Have you received results from biopsy from July 19<br>**Response:** Your biopsy was negative. Please call our office to schedule a consult to discuss your plan of care. | Logistical - tests | REAL | 36 (73%) | 13 (27%) |
| **Message:** I've had my first physical seizure this morning. It was just a facial muscle twitch and pulse no passing out.<br>**Response:** I'm sorry to hear that you had a seizure. It's really important to figure out why it happened. Let's make an appointment so we can talk more about it and decide what to do next. | Medical - problems | GPT | 35 (71%) | 14 (29%) |
| **Message:** I just read the x-ray report from my visit. I'm concerned with the Cardiomegaly. Is the enlarged heart due to the stomach being in my chest?<br>**Response:** Cardiomegaly means that the heart is bigger than it should be, and there can be different reasons for this. It's not likely to be connected to the stomach being in the chest. I suggest talking to your doctor about the results to get more information and to understand what it means. | Medical - tests | GPT | 32 (65%) | 17 (35%) |
| **Message:** Jacob's mouth sore is really bothering him to talk and eat, and kinda swollen. He is going to get the mouthwash in the morning. How will we know if he needs to see a doc or do we just treat the symptoms?<br>**Response:** If the sore in your mouth keeps bothering you or gets worse even after using mouthwash, or if you notice any signs of infection like a fever or redness that is spreading, it's a good idea to see a doctor. They can check it out and give | Medical - problems | GPT | 28 (57%) | 21 (43%) |
| you the right treatment. |  |  |  |  |
| **Message:** This thing in my throat is continuing to progress. I now have a steady dull pain along with a few random stabbing pains in that area. Swallowing has become a bigger issue also. I have an appointment with you this Friday, and with speech pathology afterwards. I am ready to get this thing out!<br>**Response:** I tried to call you several times today. I am sorry to have not gotten in touch. I hope that your symptoms do not worsen rapidly. If they do, you can come to ER here at anytime day or night and we will take care of you | Medical - problems | REAL | 11 (22%) | 38 (78%) |
| **Message:** I have an appointment with [DOCTOR] today at 1 and I will have him talk to Jane too, but I'm worried. Since late November, I have been on a mania-like rollercoaster of anxiety that is affecting my sleep. I'm just worried it's going to start physically affecting me. Is there anything that I can do?<br>**Response:** It would be best to discuss this in the office. My staff will contact you to make an appointment. | Medical - problems | REAL | 34 | 15 |
| **Message:** Can you tell me what type of test this is: Natriuretic Peptide, as it appears that I am well over the range. Thanks.<br>**Response:** The Natriuretic Peptide test is used to | Informational - tests | GPT | 45 (92%) | 4 (8%) |
| measure the levels of a hormone that is released by the heart when it is under stress. If the levels are higher than normal, it could be a sign of heart failure or other heart-related conditions. We kindly request that you schedule an appointment to discuss your test results in more detail. |  |  |  |  |
| **Message:** We have questions re: test results. Please call when possible.<br>**Response:** I'm sorry to hear that you're confused about your test results. Don't worry, I'm here to assist you. Can you please give me more information about the test and what's bothering you? | Medical -tests | GPT | 46 (94%) | 3 (6%) |
| **Message:** Good morning. I have noticed since Sunday that I has ptosis on the right side it is not worse but it is not better. I am asymptomatic otherwise, but I get tired easy. What do you think?<br>**Response:** I'm sorry to hear about your ptosis, which is a drooping of the eyelid. It's important to look into this more. I suggest making an appointment with me so we can talk about your symptoms and do a physical exam. | Medical - problems | GPT | 42 (86%) | 7 (14%) |
| **Message:** Please let me know when you get my results<br>**Response:** [DOCTOR] believes that this probably due to recent steroids for bronchitis, but you also had some elevated white blood cell count in the past | Medical - tests | REAL | 24 (49%) | 25 (51%) |
| also. It is not uncommon in our obese patient population to have elevated white blood cell (WBC) counts. Typically this elevation is not due to underlying marrow pathology but rather reflects low-grade inflammation. Please have your PCP send us some older CBC results for comparison and establish your normal range? |  |  |  |  |
| **Message:** I discovered that I have latent TB from an occupational screening . The exposure was during some construction work years ago but they will not treat me. My daughter is immune compromised due to medications she takes. My concern is that my insurance will not treat latent TB. However if I wait until I am active it is already too late and my family has been exposed. More importantly my daughter who's immune system is suppressed.<br>**Response:** [DOCTOR] would like to meet with you in clinic to discuss in more detail. Are there days and times that work well for you? | Medical - problems | REAL | 33 (67%) | 16 (33%) |

## DISCUSSION

In this study, primary care providers evaluated the quality of synthetic versus authentic patient portal message responses. The results revealed that responses generated by GPT-4 achieved statistically higher ratings in empathy and readability, with a notable trend toward statistical differences in relevance and medical accuracy compared to typical patient portal message responses. These findings not only build upon but also validate previous research by Ayers and colleagues,^4^ where a small team of healthcare professionals rated online chatbot responses as more empathetic than verified physician responses. Our study extends these findings by including a larger set of patient portal message response types and utilizing actual primary care providers to assess response quality, thus demonstrating promising results in terms of non-inferiority. More importantly, participants in our study were experienced in responding to patient portal messages, as well as experienced primary care providers, who might have been less tolerant of the untailored messages previously possible to generate before the emergence of generative AI. This aspect underscores the significance of our findings, as they reflect the responses of healthcare professionals accustomed to the nuances of patient communication and who may have higher expectations regarding message quality and relevance.

This study emphasizes the potential transformational power of AI messaging platforms in healthcare communications. It sheds light on a future in which interactions with machines are as fluid, intuitive, and fulfilling as with other humans. As AI-enabled messaging systems continue to mature and advance, with attention to message tailoring and the specific needs of patients from diverse backgrounds, chatbots and similar tools are likely to become more commonplace in medicine. Already, several studies are exploring feasibility of integrating systems such as GPT to generate high-quality responses to patient inquiries and aid clinical decisions-making across various medical specialties.^10,15,16^

Of note, crafting effective prompts entailed iterative trial and error. The potential for performance variation underscores the importance of understanding the model’s reliance on training data patterns and ensuring the relevance and quality of examples provided. Our resulting strategy and prompts are available for reference, providing valuable insights for future research and implementation endeavors in this rapidly evolving field.

## LIMITATIONS

This study is subject to several limitations that may impact its generalizability. Firstly, the sample size of both generated messages (8) and participating providers (49) is relatively small, potentially limiting the breadth of perspectives represented. Additionally, all participants were drawn from a single healthcare system, which may not fully capture the diversity of opinions regarding the value proposition for patient portal message responses or the preferred format and comprehensiveness of these responses across different healthcare settings. Furthermore, the study relied on a convenience sample of providers who may have had more time and interest to participate in the survey, introducing a potential bias in the results. As such, caution should be exercised when generalizing the findings of this study to broader populations.

The patient portal messages used to generate these synthetic responses were generated using GPT-4. At the time of this study, we were not permitted to use even HIPAA safe harbor compliant messages outside of the health system firewall. We anticipate that health systems will relax this constraint shortly, which will facilitate larger studies within a health system. Finally, our use of prompt engineering to generate responses is currently a trial-and-error process, with features of messages proposed by our research team. It will be important to better understand the desirable characteristics of patient portal message responses from the perspective of health care providers and patients.

## FUTURE WORK

Considering the limitations of our pipeline, several areas for future research and improvement emerge. Quantitative assessments are crucial to validate the significance of each step in the pipeline, offering empirical evidence to support the theoretical justifications for the architecture’s structure. The grammar editing phase requires refinement to prevent overcorrection or unintended alterations of colloquial or non-standard language, thus preserving contextual appropriateness.

Continued exploration and adaptation of the underlying model will be necessary to align with evolving understandings of response coherence and relevance. Addressing biases and inaccuracies originating from the training data is imperative to improve system performance and mitigate potential data-driven biases in generated responses. Enhancing the system’s capacity to retain context throughout extended or complex conversations can be challenging and must be monitored. Finally, refining mechanisms for gauging user literacy levels is critical to ensure that response complexity aligns more accurately with user literacy and numeracy, thereby enhancing communication effectiveness. These areas represent fruitful avenues for future research and development to advance the capabilities of our system.

The study had inadequate power to assess the importance of some covariates that might be useful for implementing this functionality at scale, including patient and primary care provider characteristics. It will be critical to ensure their efficacy considering patient preferences, healthcare settings, and regulatory requirements. Further research should be done to understand these characteristics, as well as research to address any potential ethical and liability considerations related to automating message responses. Considering these limitations, while the pipeline offers a promising approach to generating human-like responses, ongoing research and iterative refinements are crucial to enhance its efficacy and applicability in diverse real-world scenarios. By tackling these difficulties and utilizing advances in artificial intelligence, healthcare communication may develop to meet patients’ and clinicians’ ever-changing requirements and expectations.

## CONCLUSION

The findings of this study suggest that GPT-4 generated responses are feasible and acceptable to primary care providers. Despite the small sample size and single healthcare system representation, the study provides promising insights into the potential of AI-driven messaging systems to alleviate clinician burnout and enhance patient communication. As with all technological endeavors, continual evolution is paramount for addressing challenges and leveraging emerging insights from both the technological and cognitive domain.

## ETHICS DECLARATIONS

The University of Pennsylvania Human Research Protection Program, under study No. 854147, granted approval for this research project. Participant consent was not deemed necessary as the study involved secondary data analysis of patient-portal messages, sourced through a meticulously crafted pipeline. Furthermore, the protocol for this research, also approved under study No. 854147, granted approval for retrieving the initial set of patient-portal messages from a repository at Vanderbilt Medical Center, which were later used to create synthetic patient portal messages used in the study. It is important to note that the utilization of patient portal messages from Vanderbilt Medical Center were conducted in compliance with ethical guidelines. This study did not require the patient consent for using the patient portal messages retrieved from Vanderbilt Medical Center, as the data used in this study underwent a rigorous de-identification process, rendering it impossible to trace any information back to individual patients. Thus, our research respects and upholds the principles of confidentiality and anonymity, ensuring the protection of participants’ privacy rights in accordance with established ethical standards.

## Data Availability

All data produced in the present study are available upon reasonable request to the authors
All data produced in the present work are contained in the manuscript

